# Cardiovascular and Neurological Outcomes in Patients Treated with Edoxaban for Atrial Fibrillation and Characteristics in Patients with Cancer

**DOI:** 10.1101/2023.10.25.23297577

**Authors:** Kai-Hung Cheng, Hung-Pin Tu, Kai-Chun Cheng, Marielle Scherrer-Crosbie, Ting-Yuan Hsu

**Affiliations:** Preventive Medical Center, Kao-Ho Hospital, Kaohsiung, Taiwan; Department of Public Health and Environmental Medicine, School of Medicine, College of Medicine, Kaohsiung Medical University, Kaohsiung, Taiwan; Department of Ophthalmology, Kaohsiung Medical University Hospital, Kaohsiung, Taiwan; Cardiac Ultrasound Laboratory, Division of Cardiology, Hospital of the University of Pennsylvania.

**Keywords:** edoxaban, atrial fibrillation, cancer, Alzheimer‘s disease

## Abstract

**BACKGROUND:** Direct oral anticoagulants (DOACs) outperform warfarin in vascular and bleeding events in atrial fibrillation (AF) patients. Yet, effects of DOACs on congestive heart failure (CHF) and Alzheimer’s disease (AD) remain less explored.

**METHODS:** Using the Taiwan National Health Insurance Research Database, a nationwide retrospective cohort study was conducted. The study matched 5,683 non-valvular atrial fibrillation (NVAF) edoxaban patients with 11,366 warfarin patients, and 703 NVAF with cancer (NVAF-C) edoxaban patients with 1,406 warfarin patients. Vasular and non-vascular outcomes, with focuses on CHF and AD, were compared between the edoxaban and warfarin users.

**RESULTS:** Edoxaban significantly lowered adjusted hazrad ratio (aHR) of all-cause mortality, hospitalization for gastrointestinal bleeding, and CHF (0.37, 0.74, and 0.26, respectively, in NVAF; 0.39, 0.67, and 0.31, respectively, in NVAF-C, all *p* < 0.05), compared to warfarin. Edoxaban was associated with significantly lower aHRs of acute myocardial infarction, peripheral artery disease, venous thromboembolism, pulmonary embolism, and AD (0.71, 0.48, 0.55, 0.20, and 0.66, respectively; all *p* < 0.05) in NVAF patients versus warfarin. However, edoxaban had higher aHR of hospitalized bleeding (1.19, *p* = 0.002) than warfarin in NVAF patients, but not in NVAF-C patients.

**CONCLUSIONS:** Edoxaban demonstrated lowered CHF risks in both NVAF and NVAF-C patients, and reduced AD occurrence in NVAF patients versus warfarin. These findings advocate for edoxaban’s use in AF cases.

**CLINICAL PERSPECTIVE:** *What Is New?:* The study reveals that in patients with atrial fibrillation (AF), edoxaban, a direct oral anticoagulant (DOAC), demonstrates significant advantages over warfarin. Notably, edoxaban is associated with a reduced risk of congestive heart failure (CHF) and Alzheimer’s disease (AD) when compared to warfarin.

*Clinical Implications?:* These findings have important clinical implications. Edoxaban appears to be a superior anticoagulant choice for AF patients, as it lowers the risk of CHF and AD. This highlights the potential of edoxaban to improve patient outcomes and underscores its relevance for managing AF cases.

## INTRODUCTION

Atrial fibrillation (AF) affects over 37 million individuals worldwide. Its prevalence increased by 33% in the last 20 years, and projected to increase by over 60% by 2050.^1^ AF is responsible for one-sixth of all strokes,^2^ with a greater impact observed in the East Asian population.^3^

AF management in cancer patients is especially an important part of cardio-oncology field. Cancer patients face a heightened risk of AF, independent of cancer treatment, and this risk is further amplified by surgery and chemotherapy, underscoring the importance of effective anticoagulation.^4^ Low-molecular weight heparins (LMWHs) are not approved for thromboprophylaxis in patients with cancer and AF,^5^ and managing warfarin can be challenging.^5^ Direct oral anticoagulants (DOACs) are particularly attractive in cancer patients due to their rapid onset/offset action and lower bleeding risk, especially in Asian individuals.^5^ Sub-analyses of RE-LY, ROCKET AF, ENGAGE AF, and ARISTOTLE trials showed that the incidence of warfarin-associated hemorrhagic stroke was numerically higher in East Asia, underlining the need for alternative treatments.^6^ A recent nationwide retrospective cohort study from Taiwan National Health Insurance Research Database (NHIRD) revealed that DOACs were associated with a lower risk of major adverse cardiovascular events, major adverse limb events, venous thrombosis, and major bleeding compared with warfarin in patients with AF and history of cancer.^7^

In addition to the therapeutic challenges in AF patients with cancer, the causal relationship of subclinical thrombosis with cerebral thromboembolism and neurological outcomes remains unclear. Prior research has shown that edoxaban lowers the activation and reduces the expression of proteinase activated receptors (PARs).^8^ PARs belong to a class of G protein-coupled receptors involved in coagulation, hemostasis, and inflammation, and, recently recognized as important modulators of synaptic efficacy and plasticity.^9^ The PARs pathway also modulates autophagy by facilitating the production of reactive oxygen species (ROS) and beta amyloid Aβ (1-42), the main component of the amyloid plaques found in brains of people with Alzheimer’s disease (AD).^10^ Inhibition of PAR1 improves cognitive performance and alleviates synaptic plasticity impairments in a rat model of AD.^11^

With the increased use of edoxaban in Taiwan in recent years, the study comprehensively assesses the vascular, heart failure, bleeding, and neurological outcomes in Taiwanese AF patients treated with edoxaban or warfarin, considering their cancer history. We especially focused on the unexplored impact on congestive heart failure (CHF) and non-vascular neurological outcomes like Alzheimer’s disease (AD).

## METHODS

A nationwide analysis was conducted using Taiwan NHIRD, a database of catastrophic illness and medical claims. Taiwan’s National Health Insurance system provides medical coverage to all its citizens. The NHIRD contains healthcare data from over 99% of the population. To protect confidentiality, patient identification was encrypted, and authorized researchers were only allowed to link the data. By using scrambled personal identifiers, researchers were able to link files and gather socio-demographic information and longitudinal medical history.

Patients with AF (ICD-9-CM 427.31 or ICD-10-CM I48.xx) without valvular heart disease (ICD-9-CM 394.0, 394.2, 396.1, 396.8, 396.9, 746.5 + procedure codes V43.3, 35.02, 35.12, 35.20, 35.22, 35.24, 35.26, 35.28 or ICD-10-CM I05.0, I05.2, I08.0, I08.1, I08.3, I08.8, I08.9, I09.81, I34.2, Q23.2, Q23.8, Q23.9, Z95.2 + procedure codes 02QG, 02RF, 02RG, 02RH, 02RJ) were identified from Jan. 2014 to Dec. 2018. Within the non-valvular AF (NVAF) cohort, patients with co-existent cancer (ICD-9-CM 140-209.3 or ICD-10-CM C00- C97) were identified (NVAF-C). From NVAF and NVAF-C, patients receiving edoxaban or warfarin were captured. Patients using these two medications before NVAF diagnosis were excluded. Edoxaban and warfarin cohorts were matched for sex and age (1:2) using propensity score. The study was approved by the Institutional Research Ethics Committee (IRB Number: KMUHIRB-EXEMPT(II)-21090039). Informed consent was waived for secondary data analysis.

The data from the National Death Certificate Registry, which includes the death marker and date, was used to calculate overall mortality. We connected the inpatient database to track clinical events that occurred after initiating edoxaban or warfarin, including ischemic stroke/systemic embolism (ISSE), acute myocardial infarction (AMI), congestive heart failure (CHF), intracranial hemorrhage (ICH), hospitalization for gastrointestinal bleeding (GIB), other hospitalized bleeding events (OHB), peripheral artery occlusive disease (PAOD) with gangrene, venous thromboembolism (VTE), pulmonary embolism (PE), AD, and acute kidney injury (AKI). Composite efficacy events included all-cause mortality, ISSE, AMI, and ICH; composite bleeding events included GIB and OHB.

Demographic data were presented as mean ± standard deviation (SD) or median and interquartiles. Differences between the edoxaban and warfarin groups were assessed using chi-square test or Fisher’s exact test for qualitative variables, Student’s t-test for normally distributed quantitative variables, and Wilcoxon rank sum test for non-normally distributed variables.

Age and sex were used for one to two matching to alleviate potential confounders between the two groups, with unmatched patients being excluded. Clinical events were reported as adjusted hazard ratio (aHR) with 95% confidence intervals (CI), determined using a Cox proportional hazards regression model. The model was adjusted for various factors including CHA2DS2-VASc score, HAS-BLED score, medical history, and medications. Additionally, a Fine-Gray model was employed to analyze all-cause mortality as a competing outcome, presenting results as sub-distribution hazard ratio (SubHR) and adjusted SubHR after further adjustment on covariate to confirm the Cox proportional hazards regression model. Statistical analysis was conducted using SAS 9.1 software (SAS Institute, Inc., Cary, NC), with a significance level set at P < 0.05.

## RESULTS

A total of 17,050 patients with NVAF between 2014 and 2018 were included (**Figure 1**). After matching for age and gender in NVAF patients without cancer, the median age was 73 years, median CHA2DS2-VASc score was 4.5, median HAS-BLED score was 2.7, and 58% of them were male (**Table 1**). Patients treated with edoxaban had higher CHA2DS2-VASc score, more cardiovascular (CV) risk factors, chronic diseases, medications and PCI than patients treated with warfarin. Conversely, patients treated with edoxaban had less chronic kidney disease (CKD) and coronary artery bypass graft surgery than patients treated with warfarin.

**Figure 1:**
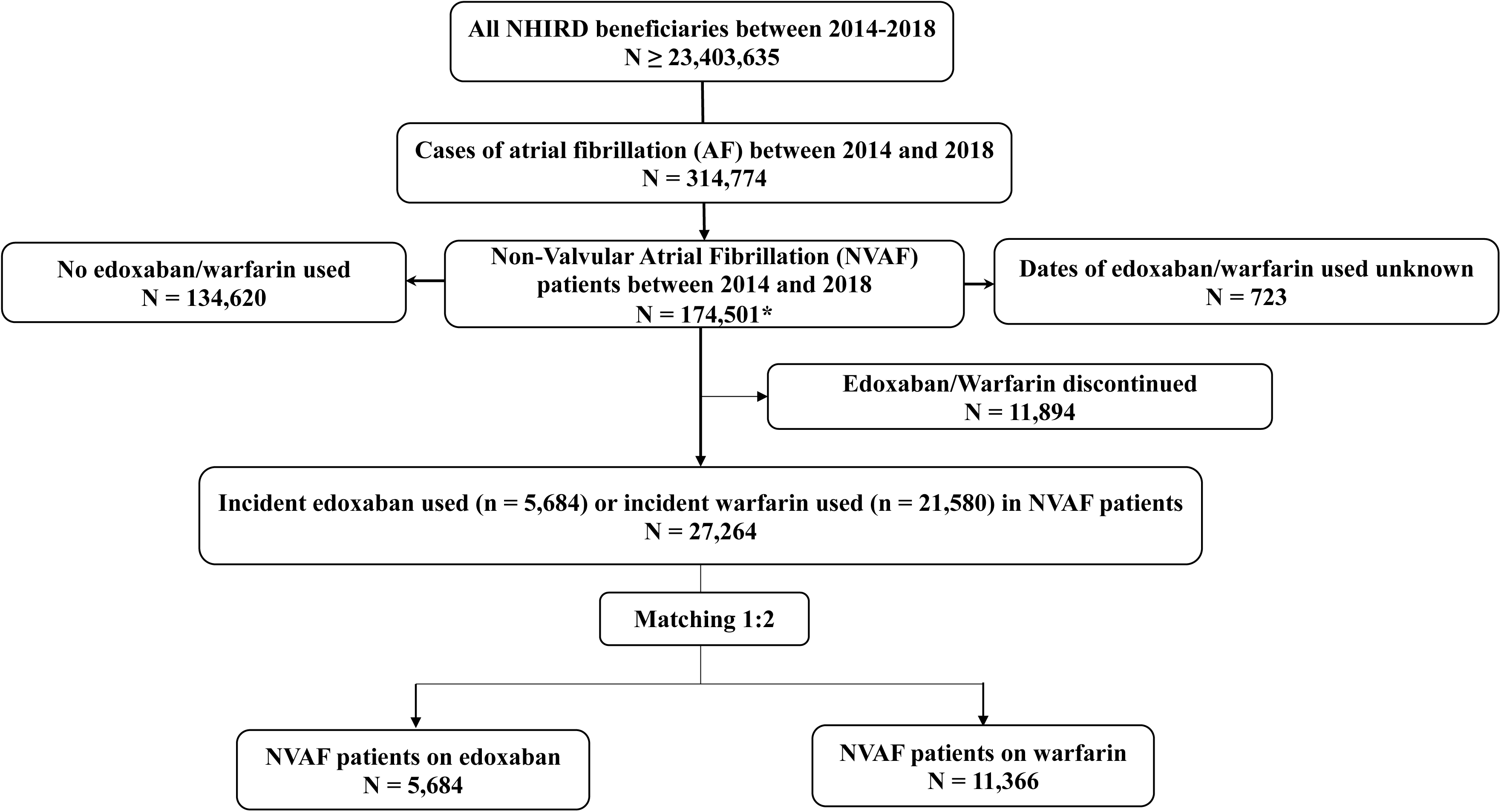
Study flow - Selection of study patients from the National Health Insurance Research Database.

**Table 1.**
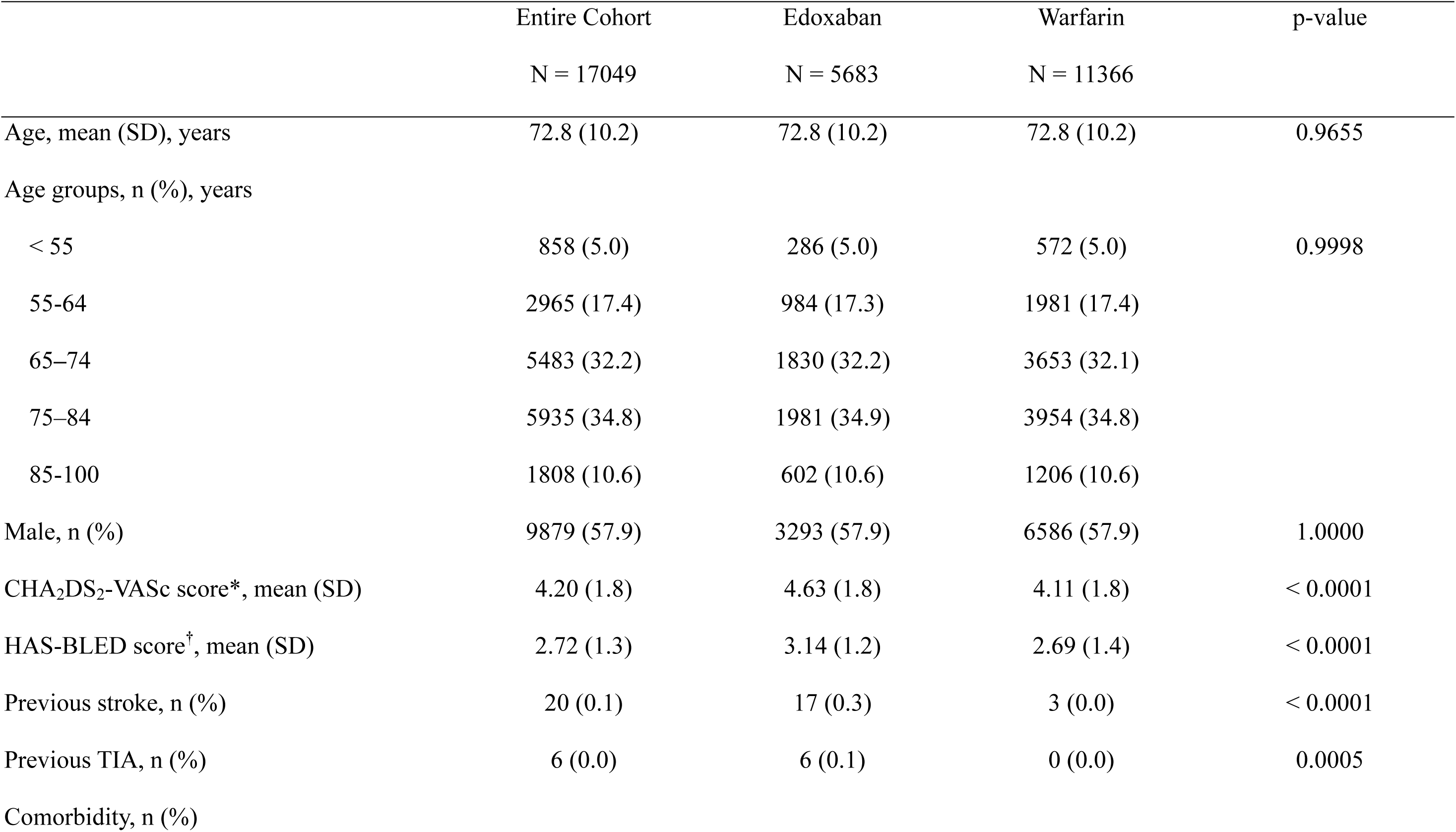
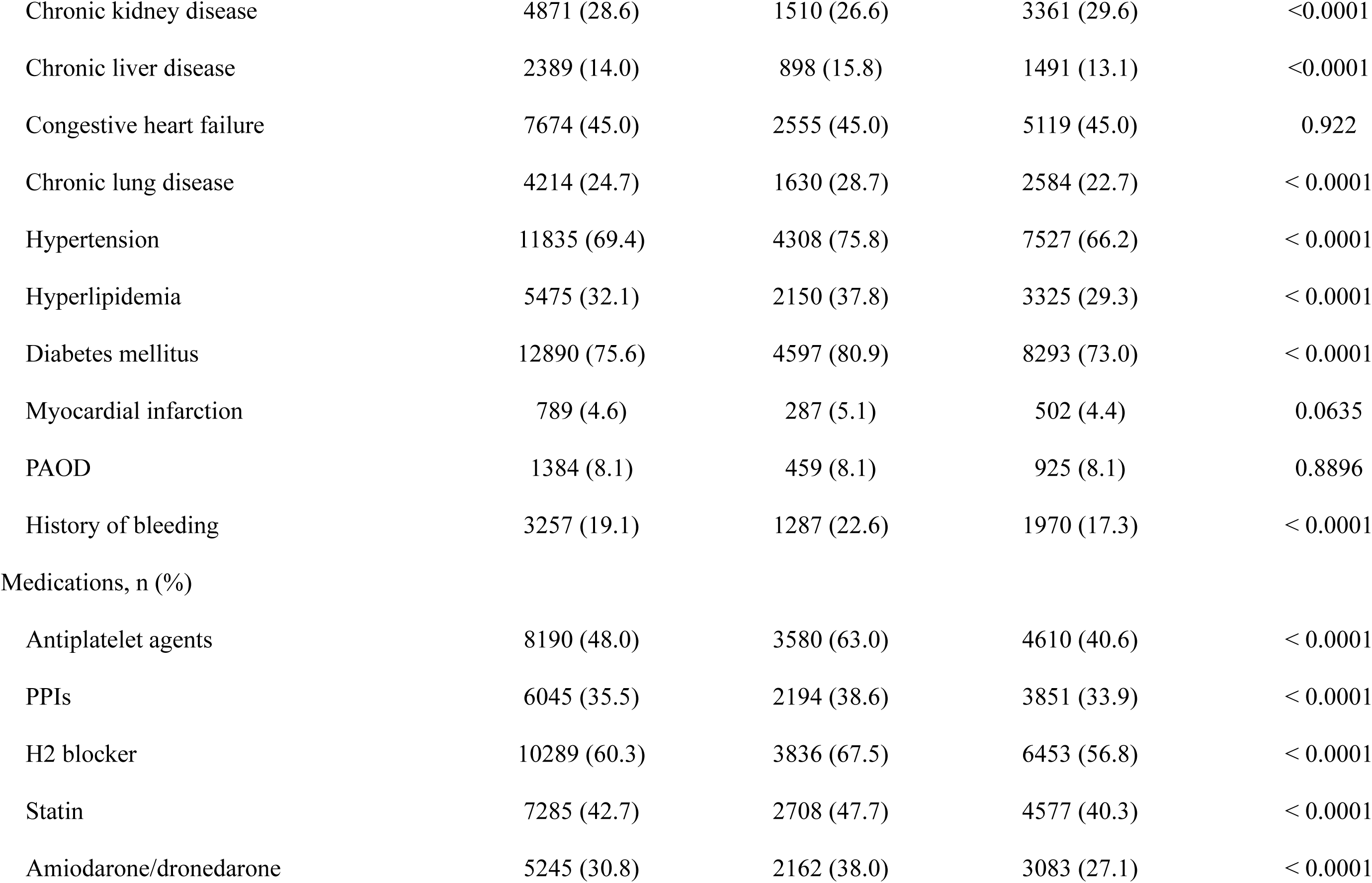
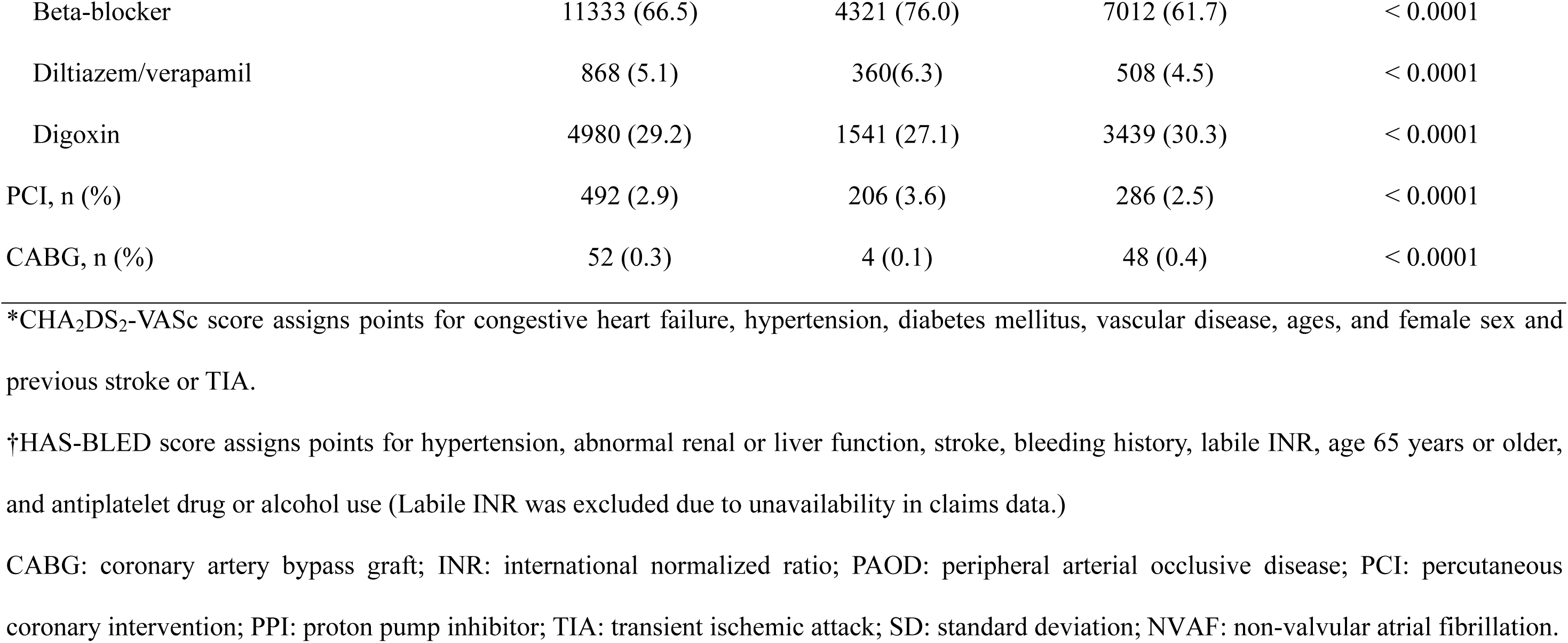
NVAF with Edoxaban vs Warfarin after 1:2 matching for sex and age.

Compared with warfarin, edoxaban was associated with significantly reduced composite efficacy events (aHR: 0.07 [0.05-0.08], *p* < 0.001), composite bleeding events (aHR: 0.13 [0.1-0.16], *p* < 0.001), all-cause mortality (aHR: 0.37 [0.33-0.42], *p* < 0.001), panarterial events (aHRs: 0.23 [0.2-0.27], *p* < 0.001), stroke, AMI, PAOD, venous thrombosis, and pulmonary embolism (**Table 2**, **Figures 2 & 3**). Edoxaban also significantly lowered aHR of CHF (0.26 [0.23-0.29], *p* < 0.001) (**Table 2**, **Figures 2 & 3-6**) versus warfarin. For neurological outcomes, edoxaban was associated with a lower aHR of AD (0.66 [0.56-0.78], *p* < 0.001) (**Table 2**, **Figure 2 & 3-10**). Fine-Gray analysis confirmed the results when all-cause mortality was modelled as a competing outcome (**Supplemental Table 1**).

**Figure 2:**
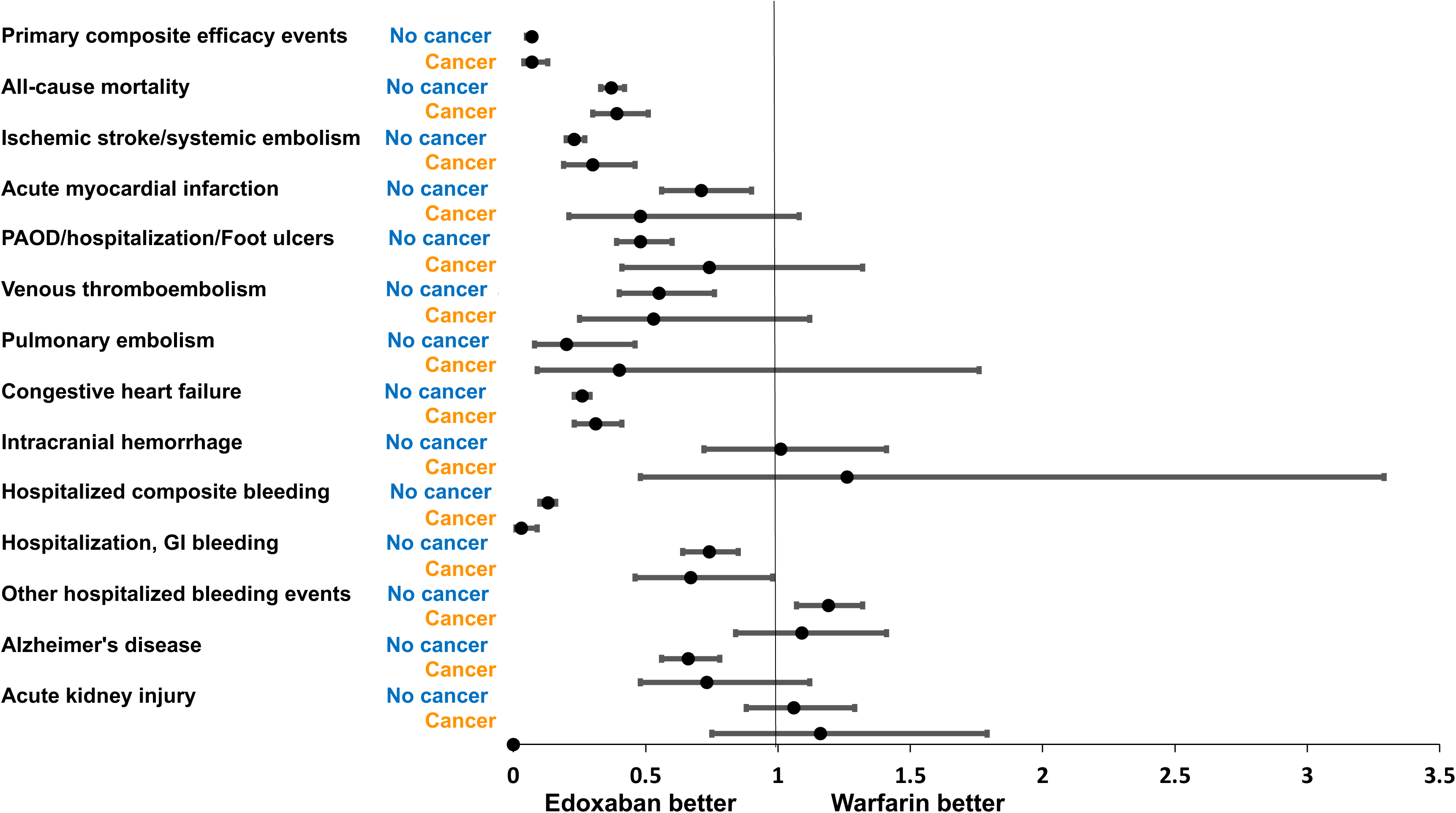
Outcomes of NVAF patients (with and without cancer) on Edoxaban or Warfarin to after 1:2 matching for sex and age.

**Figure 3:**
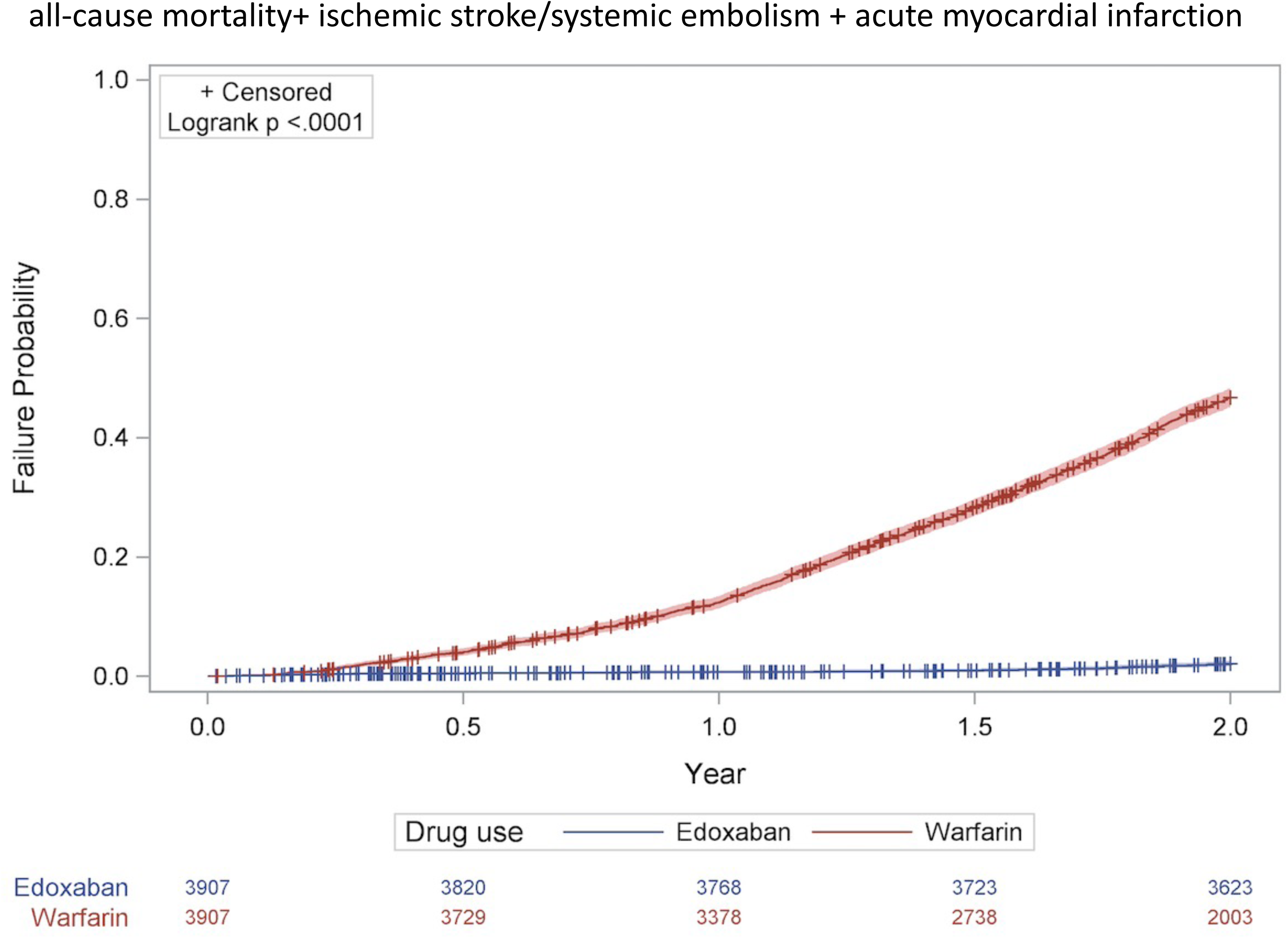
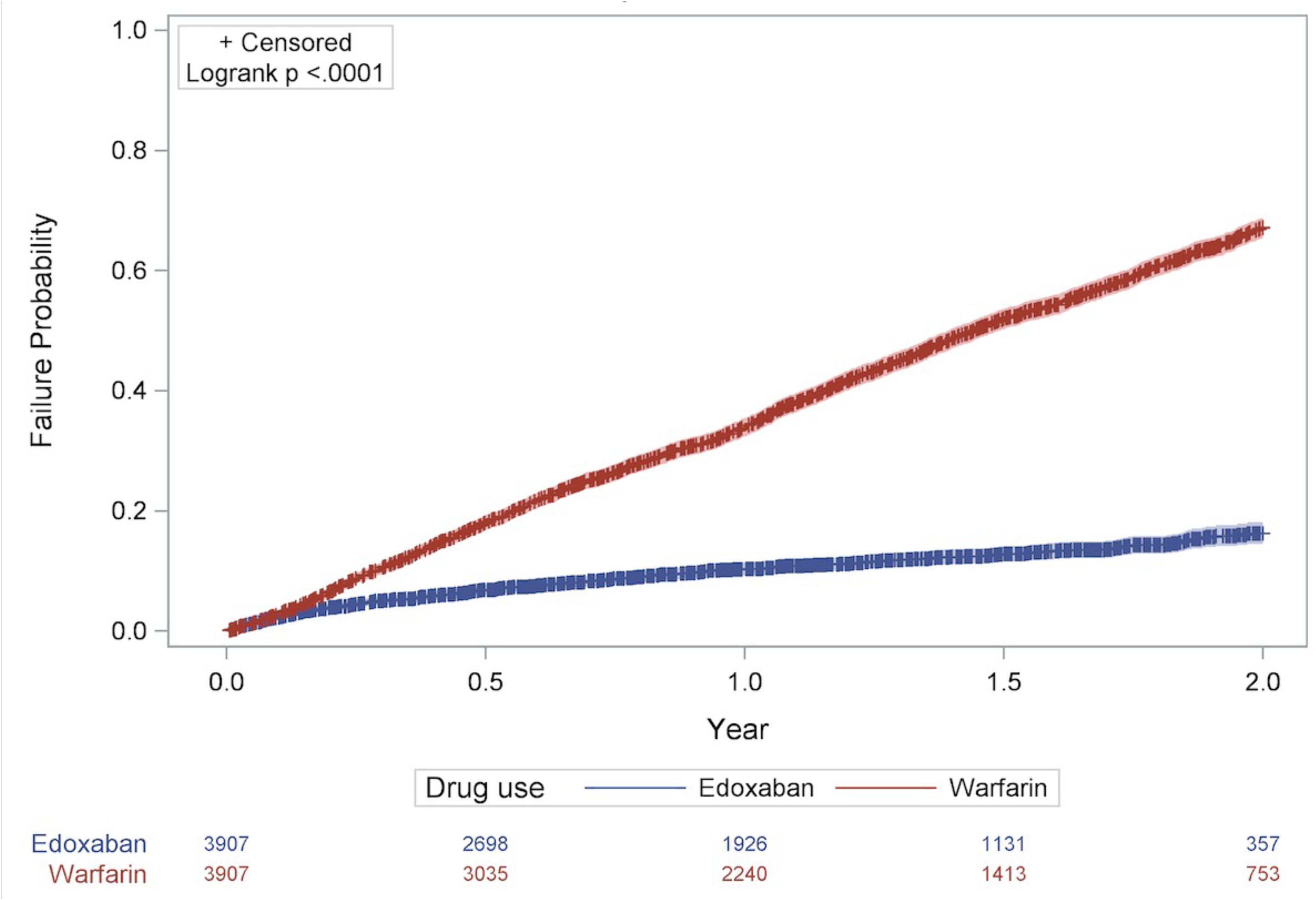
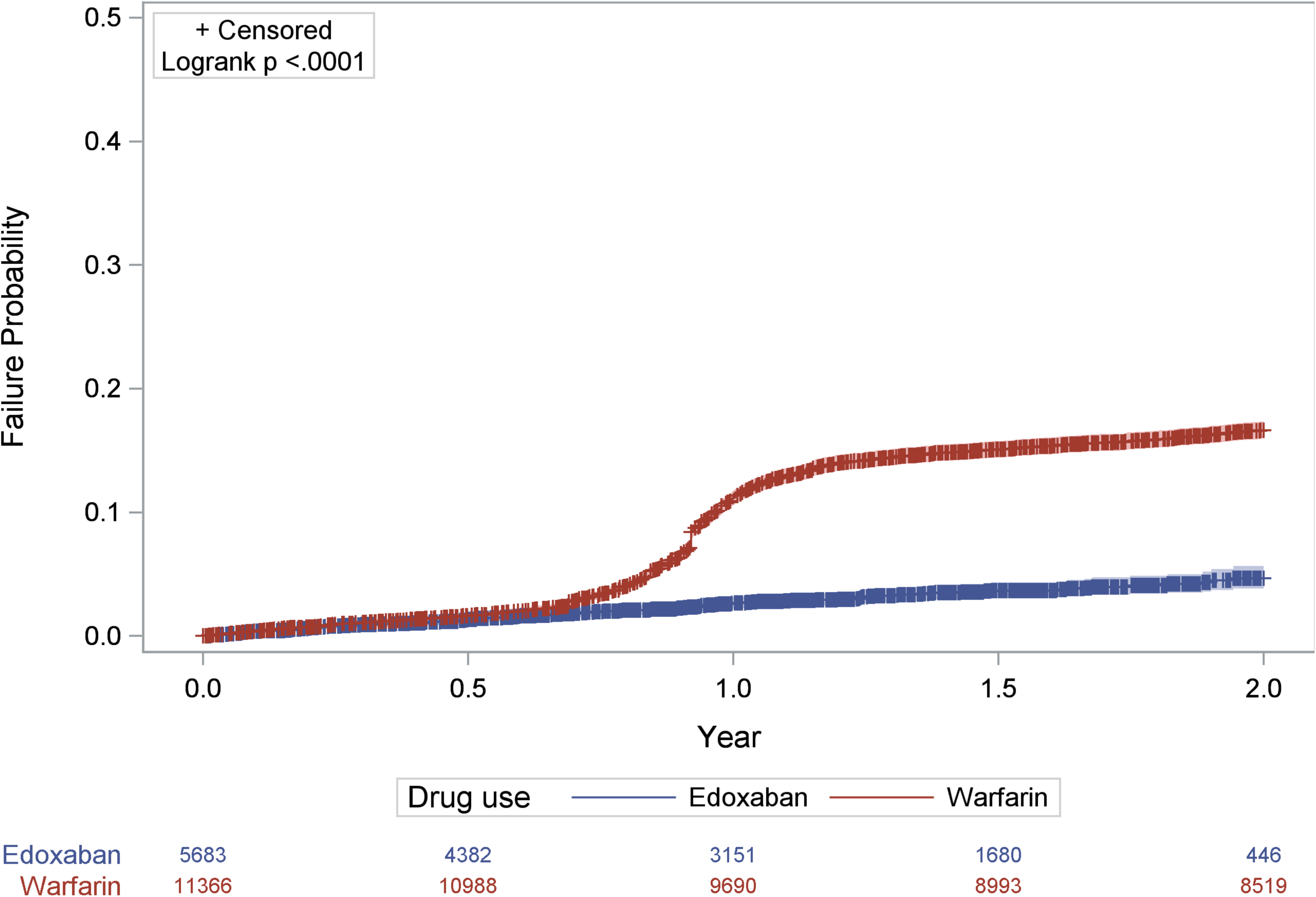
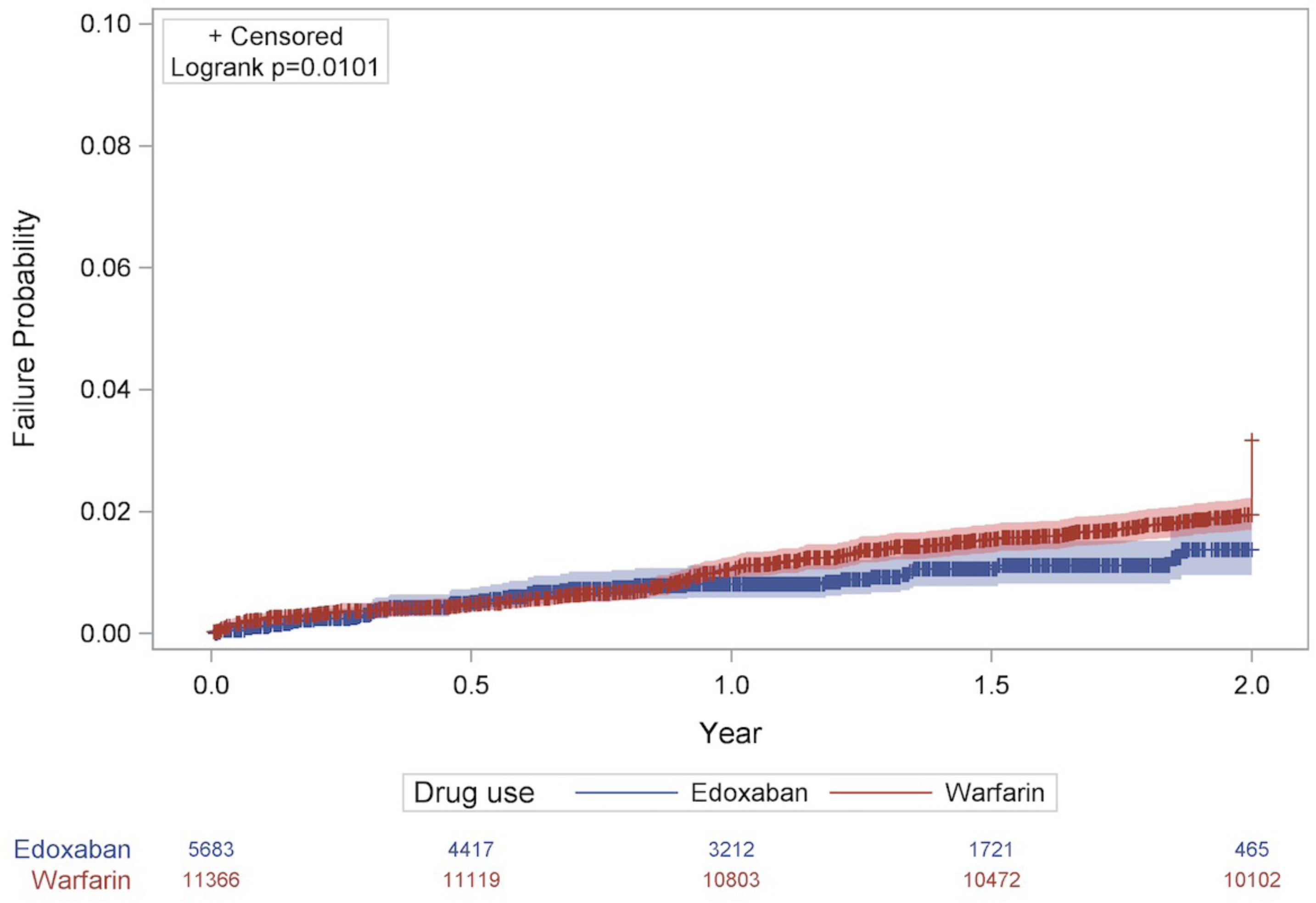
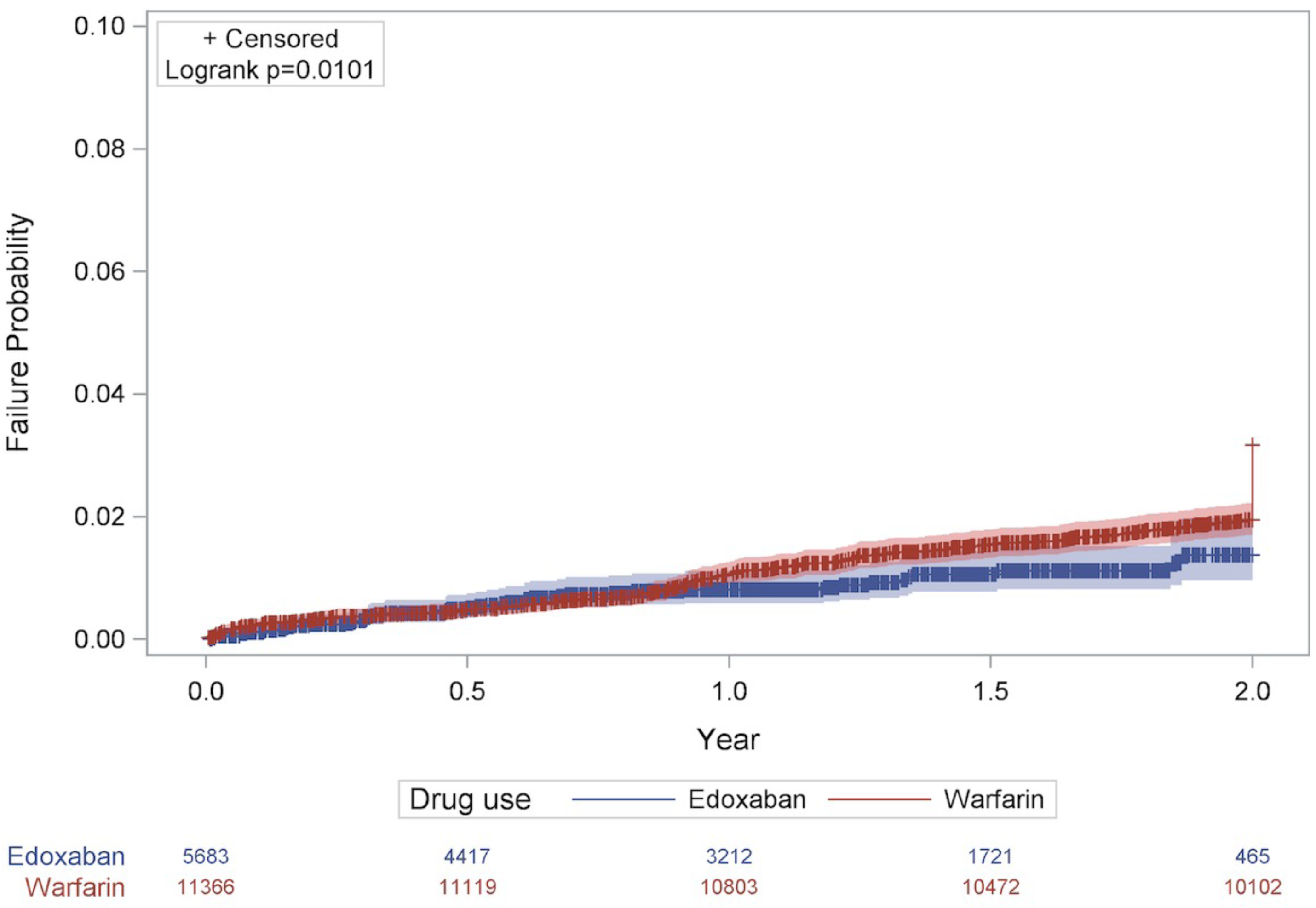
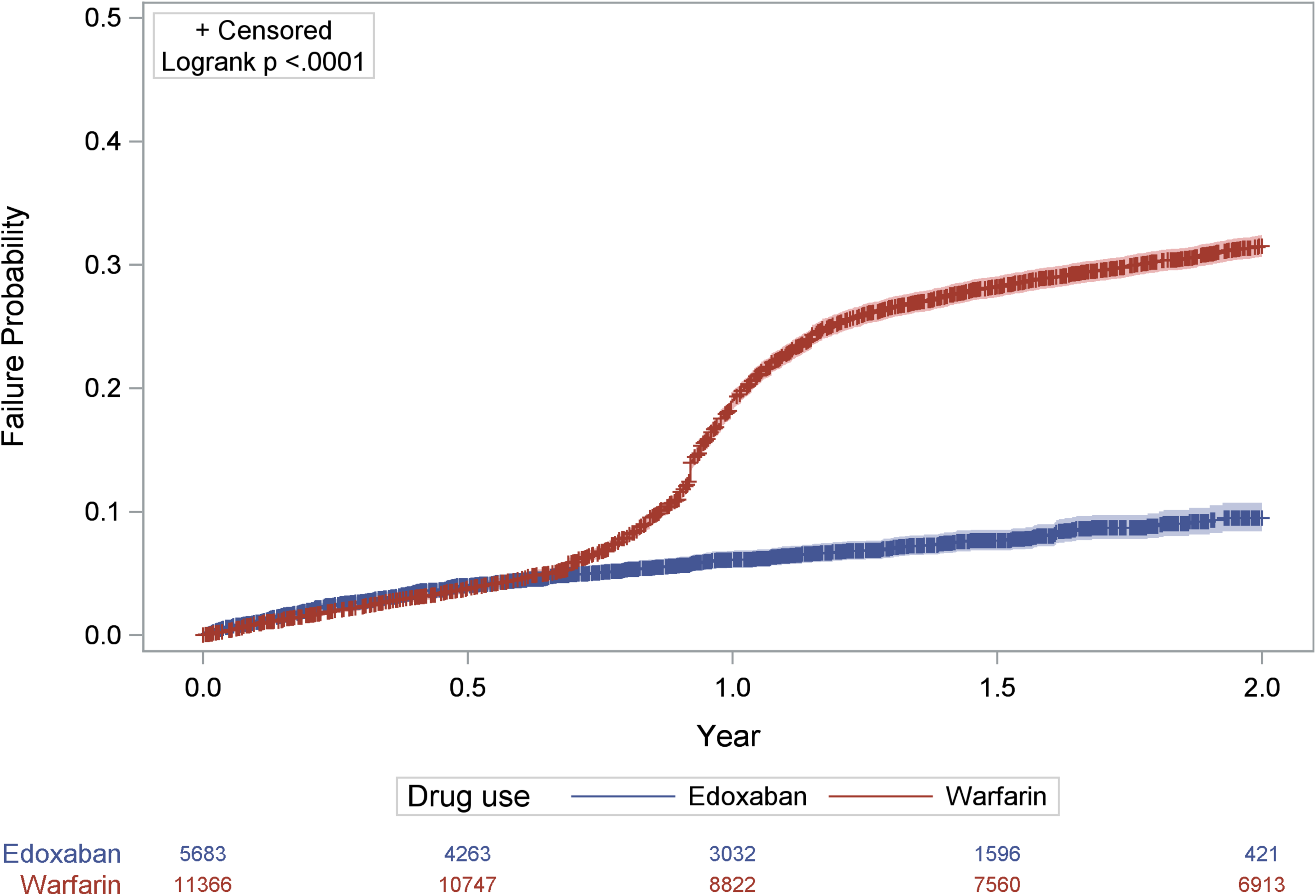
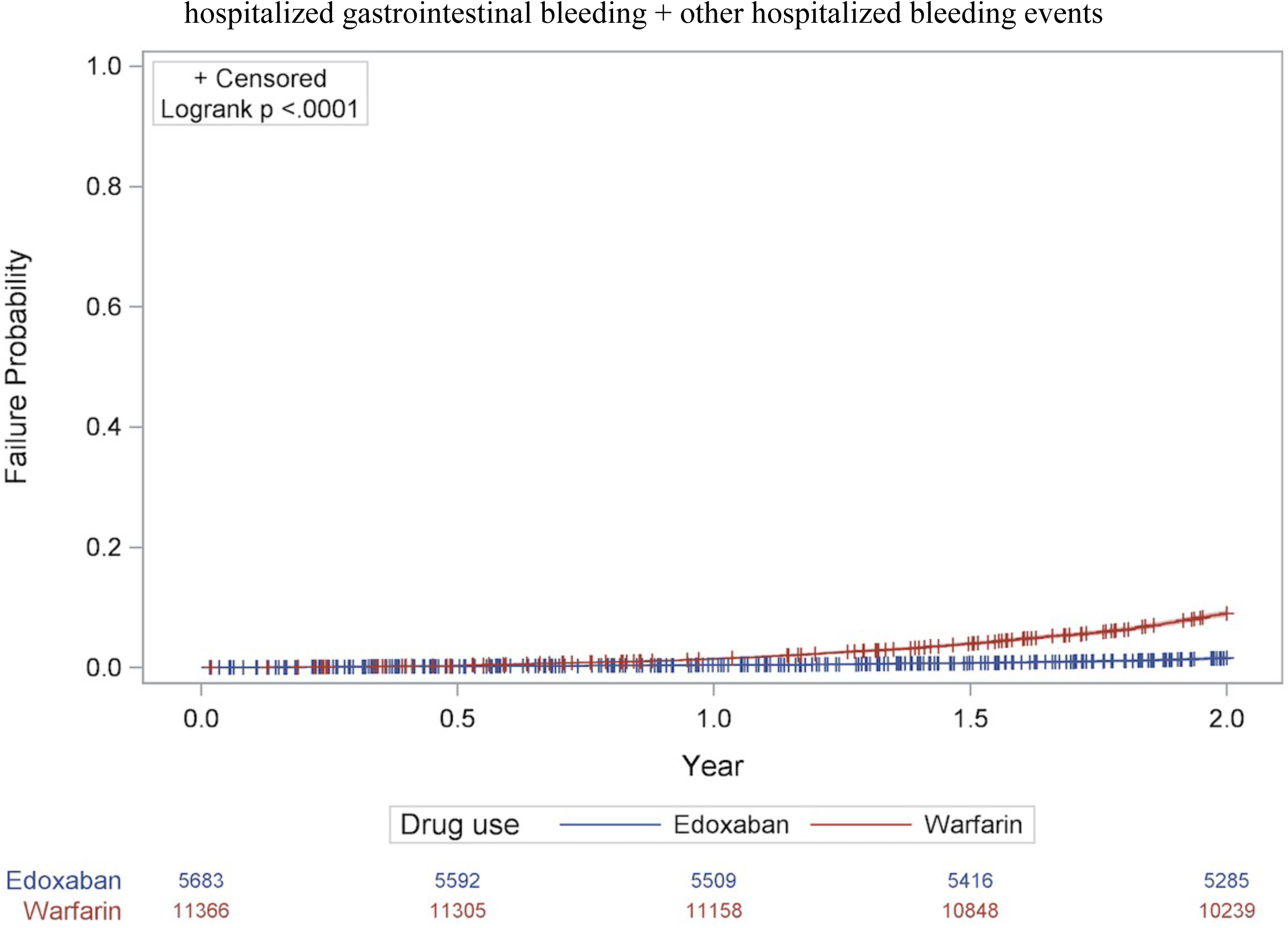
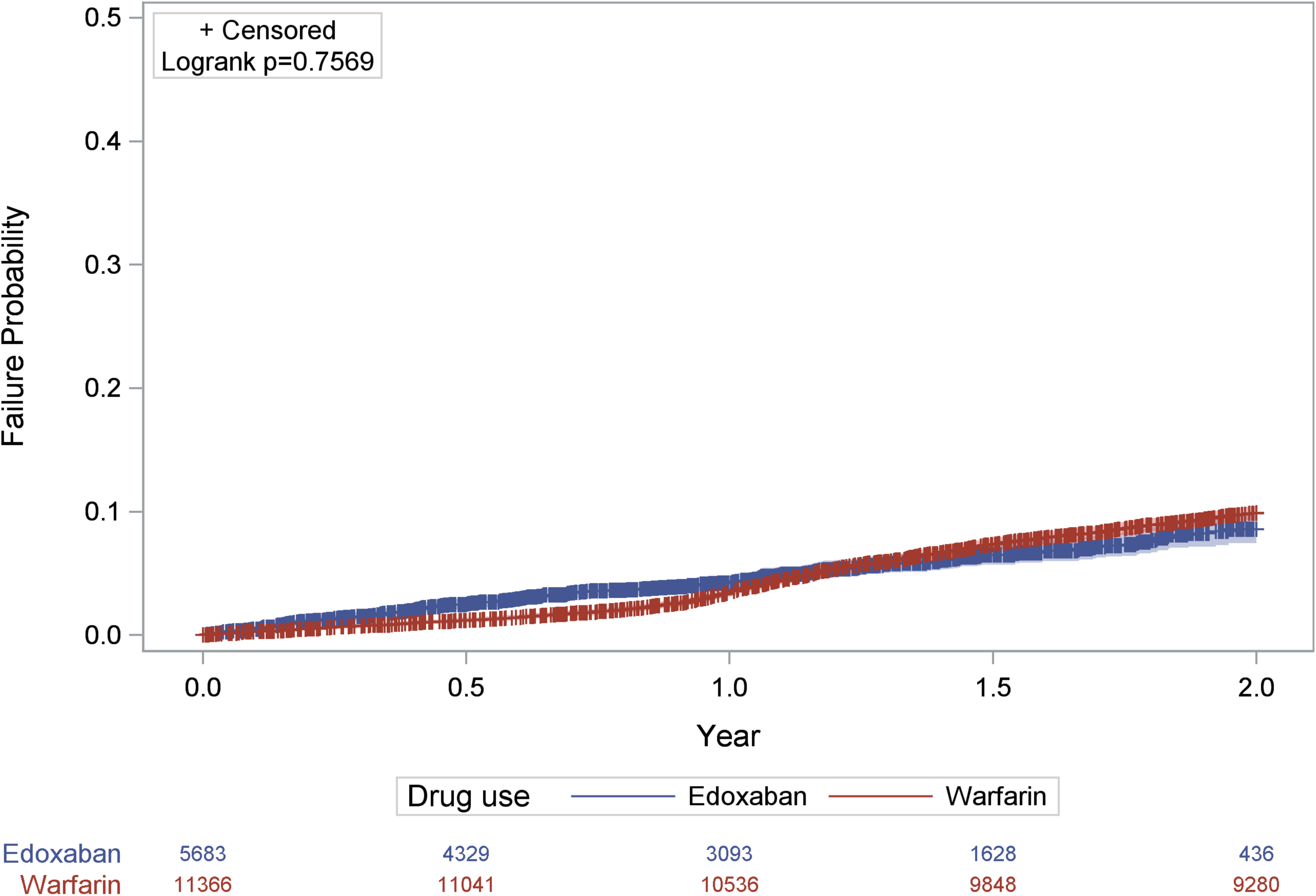
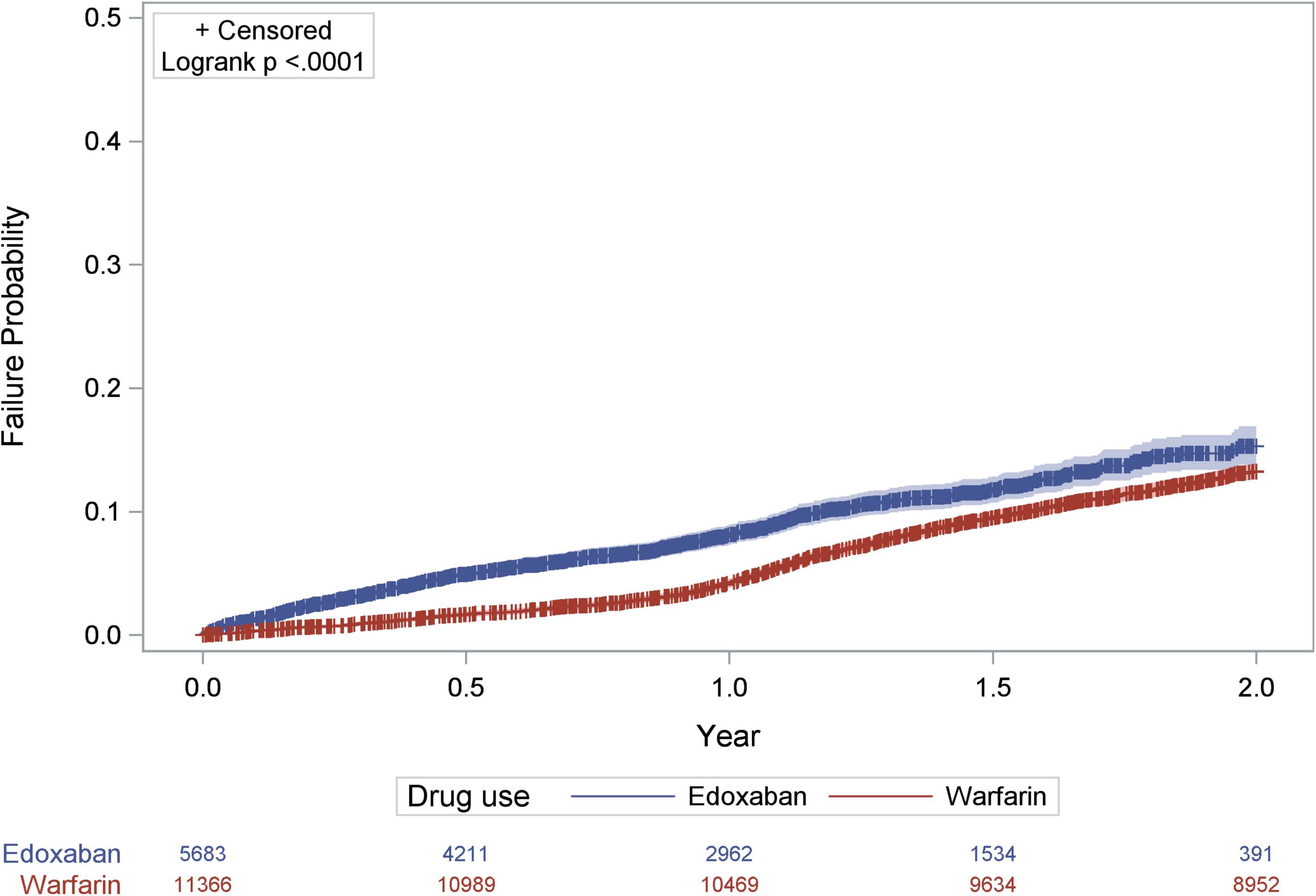
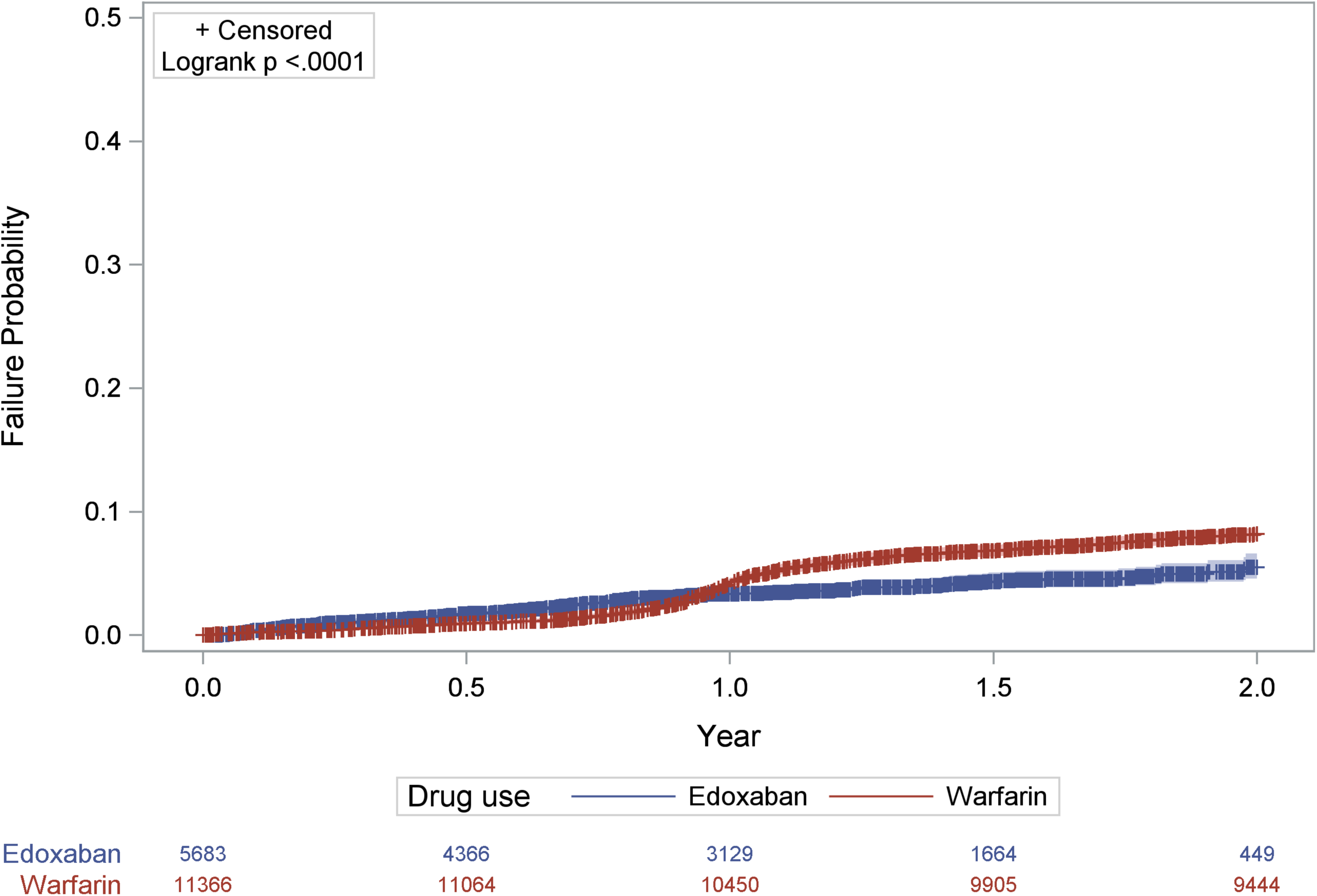
1) Primary composite efficacy events; 2) All-cause mortality; 3) Ischemic stroke/systemic embolism; 4) Overall venous thromboembolism; 5) Pulmonary embolism; 6) Congestive heart failure; 7) Hospitalized composite bleeding events; 8) Hospitalization, Gastrointestinal bleeding; 9) Other hospitalized bleeding events; 10) Alzheimer’s disease.

**Table 2.** Matched NVAF patient outcomes: Edoxaban vs. Warfarin in 2-year follow-up.

|  | Median (IQR) | Event/total, (%) | HR (95% CI) | p-value | Adjusted<br>HR (95% CI) | p-value |
| --- | --- | --- | --- | --- | --- | --- |
| Primary composite efficacy events <sup>a</sup> |  |  |  |  |  |  |
| Warfarin | 2.00 (1.35-2.00) | 1790/3907, 45.82 | 1.00 |  | 1.00 |  |
| Edoxaban | 2.00 (2.00-2.00) | 80/3907, 2.05 | 0.03 (0.03-0.04) | < 0.0001 | 0.07 (0.05-0.08) | < 0.0001 |
| All-cause mortality |  |  |  |  |  |  |
| Warfarin | 1.16 (0.56-1.83) | 2220/3907, 56.82 | 1.00 |  | 1.00 |  |
| Edoxaban | 0.98 (0.37-1.59) | 394/3907, 10.08 | 0.21 (0.19-0.23) | < 0.0001 | 0.37 (0.33-0.42) | < 0.0001 |
| Ischemic stroke/systemic embolism |  |  |  |  |  |  |
| Warfarin | 2.00 (1.99-2.00) | 1816/11366, 15.98 | 1.00 |  | 1.00 |  |
| Edoxaban | 1.11 (0.55-1.58) | 152/5683, 2.67 | 0.26 (0.22-0.31) | < 0.0001 | 0.23 (0.2-0.27) | < 0.0001 |
| Acute myocardial infarction |  |  |  |  |  |  |
| Warfarin | 2.00 (2.00-2.00) | 346/11366, 3.04 | 1.00 |  | 1.00 |  |
| Edoxaban | 1.12 (0.55-1.58) | 90/5683, 1.58 | 0.91 (0.72-1.15) | 0.4386 | 0.71 (0.56-0.9) | 0.0051 |
| PAD-gangrene/hospitalization/foot ulcers |  |  |  |  |  |  |
| Warfarin | 2.00 (2.00-2.00) | 594/11366, 5.23 | 1.00 |  | 1.00 |  |
| Edoxaban | 1.12 (0.55-1.58) | 97/5683, 1.71 | 0.55 (0.44-0.69) | < 0.0001 | 0.48 (0.39-0.60) | < 0.0001 |
| Overall venous thromboembolism |  |  |  |  |  |  |
| Warfarin | 2.00 (2.00-2.00) | 338/11366, 2.97 | 1.00 |  | 1.00 |  |
| Edoxaban | 1.13 (0.56-1.59) | 47/5683, 0.83 | 0.66 (0.48-0.91) | 0.0108 | 0.55 (0.4-0.76) | 0.0003 |
| Pulmonary embolism |  |  |  |  |  |  |
| Warfarin | 2.00 (2.00-2.00) | 87/11366, 0.77 | 1.00 |  | 1.00 |  |
| Edoxaban | 1.13 (0.56-1.59) | 6/5683, 0.11 | 0.23(0.1-0.53) | 0.0006 | 0.20 (0.08-0.46) | 0.0002 |
| Congestive heart failure |  |  |  |  |  |  |
| Warfarin | 2.00 (1.06-2.00) | 3432/11366, 30.20 | 1.00 |  | 1.00 |  |
| Edoxaban | 1.07 (0.50-1.55) | 346/5683, 6.09 | 0.30 (0.27-0.34) | < 0.0001 | 0.26 (0.23-0.29) | < 0.0001 |
| Intracranial hemorrhage |  |  |  |  |  |  |
| Warfarin | 2.00 (2.00-2.00) | 138/11366, 1.21 | 1.00 |  | 1.00 |  |
| Edoxaban | 1.13 (0.56-1.59) | 50/5683, 0.88 | 1.27 (0.91-1.77) | 0.1546 | 1.01 (0.72-1.41) | 0.9749 |
| Hospitalized composite bleeding events <sup>b</sup> |  |  |  |  |  |  |
| Warfarin | 2.00 (2.00-2.00) | 1015/11366,8.93 | 1.00 |  | 1.00 |  |
| Edoxaban | 2.00 (2.00-2.00) | 86/5683,1.51 | 0.17 (0.14-0.21) | < 0.0001 | 0.13 (0.1-0.16) | < 0.0001 |
| Hospitalization, gastrointestinal bleeding |  |  |  |  |  |  |
| Warfarin | 2.00 (2.00-2.00) | 1069/11366, 9.41 | 1.00 |  | 1.00 |  |
| Edoxaban | 1.09 (0.52-1.56) | 273/5683, 4.80 | 0.98 (0.86-1.12) | 0.7583 | 0.74 (0.64-0.85) | < 0.0001 |
| Other hospitalized bleeding events |  |  |  |  |  |  |
| Warfarin | 2.00 (2.00-2.00) | 1432/11366, 12.60 | 1.00 |  | 1.00 |  |
| Edoxaban | 1.05 (0.48-1.53) | 507/5683, 8.92 | 1.40 (1.26-1.56) | < 0.0001 | 1.19 (1.07-1.32) | 0.0016 |
| Alzheimer's disease |  |  |  |  |  |  |
| Warfarin | 2.00 (2.00-2.00) | 892/11366, 7.85 | 1.00 |  | 1.00 |  |
| Edoxaban | 1.10 (0.54-1.57) | 184/5683, 3.24 | 0.71 (0.61-0.83) | < 0.0001 | 0.66 (0.56-0.78) | < 0.0001 |
| Acute kidney injury |  |  |  |  |  |  |
| Warfarin | 2.00 (2.00-2.00) | 502/11366, 4.42 | 1.00 |  | 1.00 |  |
| Edoxaban | 1.11 (0.55-1.58) | 145/5683, 2.55 | 1.01 (0.84-1.22) | 0.9083 | 1.06 (0.88-1.29) | 0.5364 |

Adjusted HR was calculated after considering covariates, including previous stroke, previous TIA, comorbidities (chronic kidney disease, chronic liver disease, congestive heart failure, chronic lung disease, hypertension, hyperlipidemia, diabetes mellitus, myocardial infarction, PAOD), history of bleeding (The factors related to the CHA2DS2-VASc score and HAS-BLED score are already included in the information provided above), and medications.
<sup>a</sup> Primary composite efficacy events included all-cause mortality, ischemic stroke/systemic embolism, and acute myocardial infarction.
<sup>b</sup> Hospitalized composite bleeding events included hospitalized gastrointestinal bleeding and other hospitalized bleeding events, except for ICH.
Note: All-cause mortality and primary composite efficacy events were matched for medication and follow-up period due to the shorter follow-up period for edoxaban in this database.
HR: hazard ratio; IQR: interquartile range; CI: confidence of interval; PAD: peripheral arterial disease; NVAF: non-valvular atrial fibrillation

After matching for age and gender in NVAF-C patients, the mean age was 75 years, 59% of patients were male, their median CHA2DS2-VASc score was 4, and the median HAS-BLED score was 2.9. After matching for age and gender, NVAF-C patients treated with edoxaban had higher CHA2DS2VASC score, more CV risk factors, chronic diseases, medications than NVAF-C patients treated with warfarin, but less CKD than those treated with warfarin (**Table 3**).

**Table 3.** NVAF Cancer patients with Edoxaban vs Warfarin after 1:2 matching for sex and age.

|  | Entire Cohort | Edoxaban | Warfarin | p-value |
| --- | --- | --- | --- | --- |
|  | N = 2109 | N = 703 | N = 1406 |  |
| Age, mean (SD), years | 75.1 (9.0) | 75.1 (9.0) | 75.1 (8.9) | 0.9249 |
| Age groups, n (%), years |  |  |  |  |
| < 55 | 30 (1.4) | 10 (1.4) | 20 (1.4) | 1.0000 |
| 55-64 | 276 (13.1) | 92 (13.1) | 184 (13.1) |  |
| 65-74 | 678 (32.1) | 226 (32.1) | 452 (32.1) |  |
| 75-84 | 840 (39.8) | 280 (39.8) | 560 (39.8) |  |
| 85-100 | 285 (13.5) | 95 (13.5) | 190 (13.5) |  |
| Male, n (%) | 1452 (68.8) | 484 (68.8) | 968 (68.8) | 1.0000 |
| CHA<sub>2</sub>DS<sub>2</sub>-VAsC score*, mean (SD) | 4.2 (1.5) | 4.3 (1.4) | 4.1(1.5) | 0.0368 |
| HAS-BLED score<sup>†</sup>, mean (SD) | 2.9 (1.3) | 3.1 (1.2) | 2.9 (1.3) | 0.0002 |
| Previous stroke, n (%) | 1 (0.0) | 1 (0.1) | 0 (0.0) | 0.1572 |
| Previous TIA, n (%) | 1 (0.0) | 1 (0.1) | 0 (0.0) | 0.1572 |
| Comorbidity, n (%) |  |  |  |  |
| Chronic kidney disease | 752 (35.7) | 210 (29.9) | 542 (38.5) | < 0.0001 |
| Chronic liver disease | 452 (21.4) | 154 (21.9) | 298 (21.2) | 0.7075 |
| Congestive heart failure | 1006 (47.7) | 332 (47.2) | 674 (47.9) | 0.7579 |
| Chronic lung disease | 651 (30.9) | 254 (36.1) | 397 (28.2) | 0.0002 |
| Hypertension | 1607 (76.2) | 567 (80.7) | 1040 (74.0) | 0.0007 |
| Hyperlipidemia | 665 (31.5) | 231 (32.9) | 434 (30.9) | 0.3535 |
| Diabetes mellitus | 1769 (83.9) | 614 (87.3) | 1155 (82.1) | 0.0022 |
| Myocardial infarction | 96 (4.6) | 34 (4.8) | 62 (4.4) | 0.6576 |
| PAOD | 186 (8.8) | 59 (8.4) | 127 (9.0) | 0.6251 |
| History of bleeding | 455 (21.6) | 174 (24.8) | 281 (20.0) | 0.0121 |
| Medications, n (%) |  |  |  |  |
| Anthracycline | 76 (3.6) | 23 (3.3) | 53 (3.8) | 0.5631 |
| Doxorubicin | 41 (1.9) | 15 (2.1) | 26 (1.8) | 0.6555 |
| Epirubicin | 35 (1.7) | 8 (1.1) | 27 (1.9) | 0.1849 |
| Zorubicin | 1 (0.1) | 0 (0.0) | 1 (0.1) | 1.0000 |
| Idarubicin | 1 (0.1) | 0 (0.0) | 1 (0.1) | 1.0000 |
| Antiplatelet agents | 1056 (50.1) | 427 (60.7) | 629 (44.7) | < 0.0001 |
| PPIs | 1093 (51.8) | 363 (51.6) | 730 (51.9) | 0.9019 |
| H2 blocker | 1454 (68.9) | 526 (74.8) | 928 (66.0) | < 0.0001 |
| Statin | 876 (41.5) | 297 (42.2) | 579 (41.2) | 0.6393 |
| Amiodarone/dronedarone | 680 (32.2) | 269 (38.3) | 411 (29.2) | < 0.0001 |
| Beta-blocker | 1476 (70.0) | 533 (75.8) | 943 (67.1) | < 0.0001 |
| Diltiazem/verapamil | 137 (6.5) | 44 (6.3) | 93 (6.6) | 0.7548 |
| Digoxin | 651 (30.9) | 205 (29.2) | 446 (31.7) | 0.2302 |
| PCI, n (%) | 64 (3.0) | 22 (3.1) | 42 (3.0) | 0.8575 |
| CABG, n (%) | 4 (0.2) | 0 (0.0) | 4 (0.3) | 0.1569 |

\*CHA<sub>2</sub>DS<sub>2</sub>-VASc score assigns points for congestive heart failure, hypertension, diabetes mellitus, vascular disease, ages, and female sex and previous stroke or TIA.
†HAS-BLED score assigns points for hypertension, abnormal renal or liver function, stroke, bleeding history, labile INR, age 65 years or older, and antiplatelet drug or alcohol use (Labile INR was excluded due to unavailability in claims data.)
CABG: coronary artery bypass graft; INR: international normalized ratio; PAOD: peripheral arterial occlusive disease; PCI: percutaneous coronary intervention; PPI: proton pump inhibitor; TIA: transient ischemic attack; SD: standard deviation; NVAF: non-valvular atrial fibrillation.

NVAF-C patients treated with edoxaban had lower aHR of composite efficacy events (0.07 [0.04-0.13], *p* < 0.001), composite bleeding events (0.03 [0.01-0.09], *p* < 0.001), and overall mortality (0.39 [0.3-0.51], *p* < 0.001) than other treated with warfarin (**Table 4**, **Figure 2**). Additionally, aHRs for ischemic events, congestive heart failure, and GIB were lower in NVAF-C patients treated with edoxaban. The competing risk analysis confirmed the results when all-cause mortality was modelled as a competing outcome (**Supplemental Table 2)**.

**Table 4.** Matched NVAF cancer patient outcomes: Edoxaban vs. Warfarin in 2-year follow-up.

|  | Median (IQR) | Event/total, (%) | HR (95% CI) | p-value | Adjusted<br>HR (95% CI) | p-value |
| --- | --- | --- | --- | --- | --- | --- |
| Primary composite efficacy events <sup>a</sup> |  |  |  |  |  |  |
| Warfarin | 2.00 (1.77-2.00) | 172/489, 35.17 | 1.00 |  | 1.00 |  |
| Edoxaban | 2.00 (2.00-2.00) | 11/489, 2.25 | 0.05 (0.03-0.1) | < 0.0001 | 0.07 (0.04-0.13) | < 0.0001 |
| All-cause mortality |  |  |  |  |  |  |
| Warfarin | 1.43 (0.89-1.90) | 289/489, 59.1 | 1.00 |  | 1.00 |  |
| Edoxaban | 1.22 (0.64-1.72) | 77/489, 15.75 | 0.32 (0.25-0.41) | < 0.0001 | 0.39 (0.3-0.51) | < 0.0001 |
| Ischemic stroke/systemic embolism |  |  |  |  |  |  |
| Warfarin | 2.00 (2.00-2.00) | 229/1406, 16.29 | 1.00 |  | 1.00 |  |
| Edoxaban | 1.16 (0.57-1.64) | 23/703, 3.27 | 0.30 (0.20-0.47) | < 0.0001 | 0.30 (0.19-0.46) | < 0.0001 |
| Acute myocardial infarction |  |  |  |  |  |  |
| Warfarin | 2.00 (2.00-2.00) | 46/1406, 3.27 | 1.00 |  | 1.00 |  |
| Edoxaban | 1.18 (0.57-1.65) | 10/703, 1.42 | 0.76 (0.38-1.52) | 0.4431 | 0.48 (0.21-1.08) | 0.0773 |
| PAD-gangrene/hospitalization/foot ulcers |  |  |  |  |  |  |
| Warfarin | 2.00 (2.00-2.00) | 76/1406, 5.41 | 1.00 |  | 1.00 |  |
| Edoxaban | 1.17 (0.55-1.66) | 16/703, 2.28 | 0.65 (0.38-1.13) | 0.1260 | 0.74 (0.41-1.32) | 0.3016 |
| Overall venous thromboembolism |  |  |  |  |  |  |
| Warfarin | 2.00 (2.00-2.00) | 63/1406, 4.48 | 1.00 |  | 1.00 |  |
| Edoxaban | 1.18 (0.59-1.66) | 8/703, 1.14 | 0.53(0.25-1.12) | 0.1009 | 0.53 (0.25-1.12) | 0.1009 |
| Pulmonary embolism |  |  |  |  |  |  |
| Warfarin | 2.00 (2.00-2.00) | 16/1406, 1.14 | 1.00 |  | 1.00 |  |
| Edoxaban | 1.18 (0.59-1.66) | 2/703, 0.28 | 0.4(0.09-1.76) | 0.2870 | 0.4 (0.09-1.76) | 0.2870 |
| Congestive heart failure |  |  |  |  |  |  |
| Warfarin | 2.00 (1.09-2.00) | 431/1406, 30.65 | 1.00 |  | 1.00 |  |
| Edoxaban | 1.10 (0.53-1.59) | 53/703, 7.54 | 0.35 (0.27-0.47) | < 0.0001 | 0.31 (0.23-0.41) | < 0.0001 |
| Intracranial hemorrhage |  |  |  |  |  |  |
| Warfarin | 2.00 (2.00-2.00) | 14/1406, 1.00 | 1.00 |  | 1.00 |  |
| Edoxaban | 1.19 (0.57-1.66) | 7/703, 1.00 | 1.67 (0.66-4.23) | 0.2822 | 1.26 (0.48-3.29) | 0.6417 |
| Hospitalized composite bleeding events <sup>b</sup> |  |  |  |  |  |  |
| Warfarin | 2.00 (2.00-2.00) | 195/1406, 13.87 | 1.00 |  | 1.00 |  |
| Edoxaban | 2.00 (2.00-2.00) | 4/703, 0.57 | 0.04 (0.01-0.1) | < 0.0001 | 0.03 (0.01-0.09) | < 0.0001 |
| Hospitalization, gastrointestinal bleeding |  |  |  |  |  |  |
| Warfarin | 2.00 (2.00-2.00) | 166/1406, 11.81 | 1.00 |  | 1.00 |  |
| Edoxaban | 1.15 (0.55-1.64) | 38/703, 5.41 | 0.84 (0.58-1.19) | 0.3246 | 0.67 (0.46-0.98) | 0.0384 |
| Other hospitalized bleeding events |  |  |  |  |  |  |
| Warfarin | 2.00 (1.83-2.00) | 273/1406, 19.42 | 1.00 |  | 1.00 |  |
| Edoxaban | 1.07 (0.48-1.55) | 83/703, 11.81 | 1.13 (0.88-1.45) | 0.3435 | 1.09 (0.84-1.41) | 0.5193 |
| Alzheimer's disease |  |  |  |  |  |  |
| Warfarin | 2.00 (2.00-2.00) | 137/1406, 9.74 | 1.00 |  | 1.00 |  |
| Edoxaban | 1.16 (0.57-1.64) | 28/703, 3.98 | 0.68 (0.45-1.02) | 0.0642 | 0.73 (0.48-1.12) | 0.1496 |
| Acute kidney injury |  |  |  |  |  |  |
| Warfarin | 2.00 (2.00-2.00) | 98/1406, 6.97 | 1.00 |  | 1.00 |  |
| Edoxaban | 1.16 (0.57-1.65) | 29/703, 4.13 | 0.98 (0.64-1.49) | 0.9132 | 1.16 (0.75-1.79) | 0.4954 |

Adjusted HR was calculated after considering covariates, including previous stroke, previous TIA, comorbidities (chronic kidney disease, chronic liver disease, congestive heart failure, chronic lung disease, hypertension, hyperlipidemia, diabetes mellitus, myocardial infarction, PAOD), history of bleeding (The factors related to the CHA2DS2-VASc score and HAS-BLED score are already included in the information provided above), and medications.
<sup>a</sup> Primary composite efficacy events included all-cause mortality, ischemic stroke/systemic embolism, and acute myocardial infarction.
<sup>b</sup> Hospitalized composite bleeding events included hospitalized gastrointestinal bleeding and other hospitalized bleeding events, except for ICH. Note: All-cause mortality and primary composite efficacy events were matched for medication and follow-up period due to the shorter follow-up
period for edoxaban in this database.
HR: hazard ratio; IQR: interquartile range; CI: confidence of interval; PAD: peripheral arterial disease; NVAF: non-valvular atrial fibrillation

## DISCUSSION

In this study, edoxaban demonstrated greater beneficial effects than warfarin in NVAF patients with and without cancer, based on a lower aHR of thrombotic events, CHF, and mortality. Edoxaban was also associated with significantly reduced risks of AD versus warfarin in NVAF patients without cancer. Besides, edoxaban showed a significantly better safety profile than warfarin in composite bleeding events in NVAF patients with and without cancer. Surprisingly, this study revealed some noteworthy findings regarding impacts of DOACs on AD and CHF that had not been previously discussed.

Besides anticoagulation properties, emerging data highlight additional benefits of DOACs through PARs inhibition. FXa, both *in vitro and in vivo* studies, has been found to promote atherosclerosis through PARs pathways^12^ and animal studies demonstrated that FXa inhibition reduced atherosclerosis, promoted plaque regression, reduced macrophage infiltration, enhanced collagen deposition, and decreased necrotic core size through PAR-1 and PAR-2 inhibition.^12^ Rivaroxaban also inhibited atherogenesis by suppressing macrophage autophagy and inflammasome activity mediated by FXa-PAR2.^13^ DOACs inhibited FXa and had anti-inflammatory effects, which reduced the progression of cardiac valve calcification and deterioration of renal function.^14^ In contrast, warfarin inhibited matrix gamma-carboxyglutamate Gla protein (MGP), promoting systemic calcification in the coronary and peripheral vasculature, valve, and heart.^15^ Moreover, a higher burden of coronary artery calcium was found to be closely linked to a higher risk of sudden cardiac death.^16^ These mechanisms may contribute to the lower incidence of thrombotic events and the higher survival associated with edoxaban compared to warfarin.

Our study is the first to report that edoxaban is linked to a lower rate of CHF admissions compared to warfarin in patients with NVAF, both with and without cancer. AF frequently coexists with CHF, and the joint occurrence of AF and CHF may further worsen prognosis.^17^ There are some plausible explanations for edoxaban’s benefits on CHF. First, edoxaban effectively reduced AMI events, a crucial risk factor for CHF.^18,19^ Furthermore, the renin-angiotensin-aldosterone system (RAAS) is essential in the pathophysiology of heart failure.^20^ The blockade of PAR-1 signaling, such as that induced by DOAC, was recently shown to reverse RAAS activation and attenuate cardiac fibrosis and hypertrophy.^19^ PAR-2 signaling, which can be directly activated by FXa, also contributes to the pathogenesis of hypertrophy and CHF.^21^ Finally, edoxaban decreases the incidence of AKI, which may contribute to CHF. Therefore, edoxaban might significantly reduce CHF through reduction of AMI and AKI, and by inhibiting PARs.

Our results showed that edoxaban, compared to warfarin, significantly lowered risks of AD in patients with NVAF, which was in line with previous studies. A recent review reported potential positive effects of anticoagulant on vascular component of AD and dementia.^22^ Studies in mice models also showed that normalizing the procoagulant state in AD led to a reduction in AD pathological features, suggesting the impact of coagulation on AD pathogenesis.^23^ Furthermore, DOACs diminish oxidative stress, neuroinflammation and blood–brain barrier dysfunction, which, in turn, ameliorate memory loss, and the amyloid load in AD mouse models.^24,25^ Therefore, edoxaban, as an DOAC, may prevent dementia by a direct effect on the development of AD, and by protective effects on vascular and embolic brain impairment.

An association between AF and AD has been reported, with an increase in risks of AD by 1.5-2.5 folds in AF patients.^26^ AF patients exhibit elevated thrombin level which can activate PAR-1 and cause nerve fiber conduction block.^27^ Moreover, PARs influence autophagy by affecting ROS production and the degradation of beta amyloid Aβ (1-42).^10^ Edoxaban may ameliorate cognitive deterioration of AD^11^ by reducing the expression of PARs, proinflammatory and profibrotic genes.^28^

Autopsy studies found small strokes in AD patients^19,29^ and microemboli from cardiac and vascular sources can cause brain structural changes.^29^ A recent study showed that in AF patients, even 89.9% were anticoagulated, 5.5% had a new brain infarct on MRI after 2 years and most infarcts were clinically silent.^30^ Both overt and silent brain infarcts led to cognitive decline.^30^ Therefore, differentiating vascular dementia from AD is challenging, leading to potential inclusion of vascular dementia cases within AD.

In a previous study, comparing DOACs with warfarin in AF patients with cancer, apixaban showed lower risks of ISSE and major bleeding, while dabigatran and rivaroxaban showed similar risks. ^31^ Our study provides new insights that edoxaban significantly reduced ISSE and GIB in both NVAF patients with and without cancer.

Cancer is a heterogeneous disease; therefore, results should be interpreted with caution. However, we still observed significantly greater clinical benefits on stroke and MI prevention for edoxaban over warfarin, with preserved safety profile for edoxaban. In evaluating risks and benefits, edoxaban seemed to be preferable to warfarin for NVAF patients with cancer.

There are several limitations. First, the study was retrospective and could be biased by confounding factors; therefore, we used propensity score matching to balance baseline characteristics between two groups. Second, the study was conducted in Taiwan, and results may not be generalizable to other populations with different races and healthcare systems. Third, the study did not adjust confounding factors such as smoking status and body mass index, which may have influenced outcomes. Lastly, the follow-up period was relatively short (up to 2 years), and long-term outcomes was not fully captured.

In Taiwanese patients with NVAF, edoxaban, as compared with warfarin, was associated with reduced CHF in both NVAF patients with and without cancer, and lower risks of AD in NVAF patients without cancer. All-cause mortality, ISSE, and GIB events were also significantly reduced with edoxaban, along with preserved safety profile as reflected by lower composite bleeding events in both NVAF patients with and without cancer.

## Nonstandard Abbreviations and Acronyms

AF: Atrial Fibrillation
CHF: Congestive Heart Failure
AD: Alzheimer’s Disease
DOACs: Direct Oral Anticoagulants
NVAF: Non-Valvular Atrial Fibrillation
NHIRD: National Health Insurance Research Database
LMWHs: Low-Molecular-Weight Heparins
FXa: Factor Xa
PARs: Proteinase Activated Receptors
RAAS: Renin-Angiotensin-Aldosterone System
AMI: Acute Myocardial Infarction
PCI: Percutaneous Coronary Intervention
CKD: Chronic Kidney Disease
VTE: Venous Thromboembolism
PE: Pulmonary Embolism
AKI: Acute Kidney Injury
ICD-9-CM: International Classification of Diseases, 9th Revision, Clinical Modification
ICD-10-CM: International Classification of Diseases, 10th Revision, Clinical Modification

## Acknowledgements

Authors thank Daiichi Sankyo Taiwan Ltd for the financial support and assistance in publication.

## Data availability & Supplementary material

Data are available in the article and its online Supplementary material. Hung-Pin Tu had full access to all the data in the study and takes responsibility for its integrity and the data analysis. The deidentified participant data will not be shared.

## Declaration of Helsinki

The study complied with the Declaration of Helsinki, with ethical approval from the Institutional Research Ethics Committee of Kaohsiung Medical University Hospital (IRB Number: KMUHIRB-EXEMPT(II)-21090039).

---

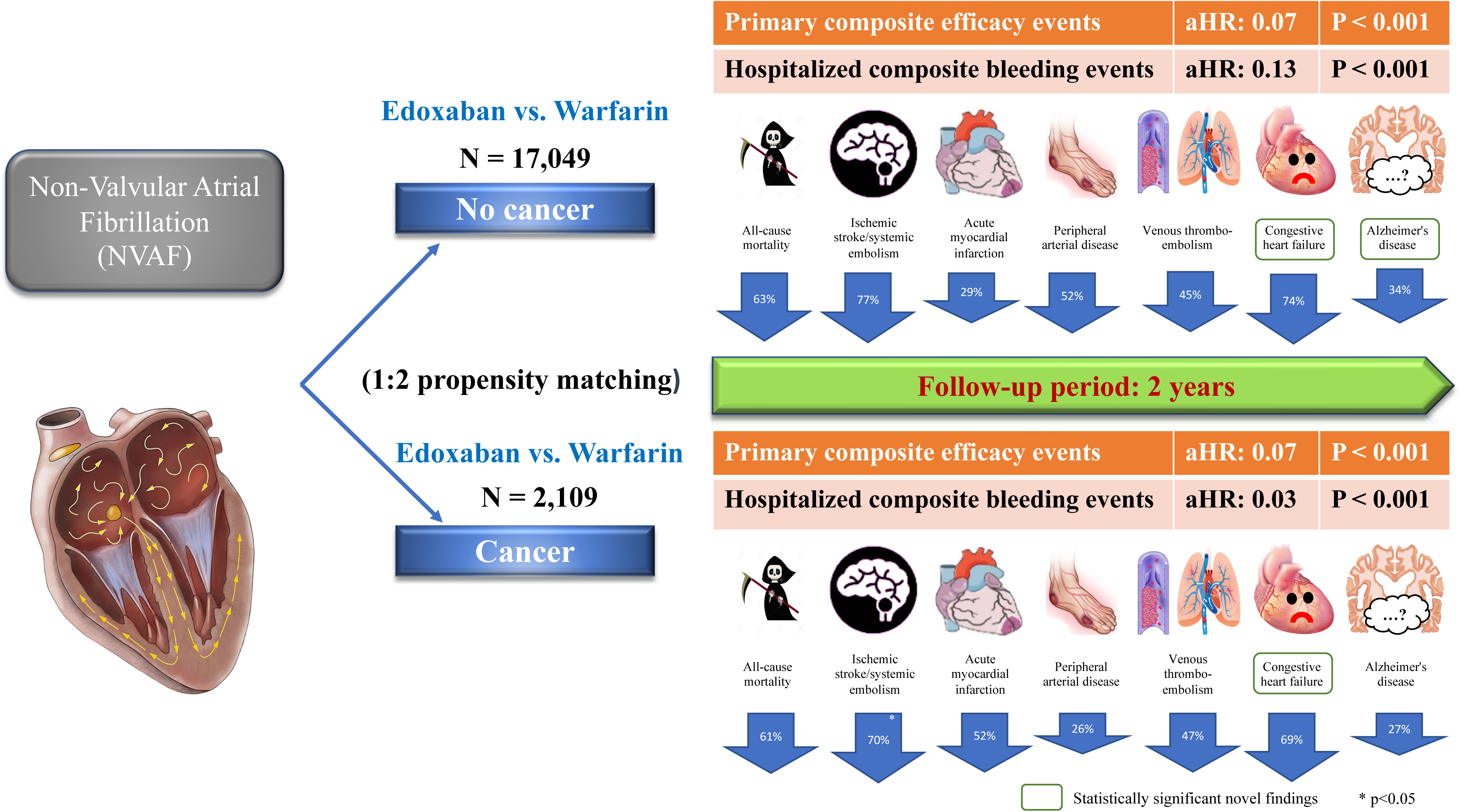

